# Multisystem inflammatory syndrome in children (MIS-C) temporally associated with SARS-CoV-2 infection: a scoping review of the literature

**DOI:** 10.1101/2020.08.03.20167361

**Authors:** Mohamed Sabbour, Seif Tarek El-Swaify, Nourhan Farrag, Menna Kamel, Sara H. Ali, Abdelrahman Amir, Mazen A. Refaat, Menatalla A. Dyab, Ashraf Nabhan

## Abstract

**Background:** With the rise of the COVID-19 pandemic, a new severe life-threatening inflammatory syndrome has been reported in some pediatric populations. Global attention was shifted towards the syndrome termed multisystem inflammatory syndrome in children (MIS-C), with new case reports flooding in.

**Objectives:** The aim of this scoping review is to summarize the existing reports on MIS-C and focus on the demographics, diagnosis, clinical presentation, laboratory investigations, imaging studies, treatment, and patient outcomes.

**Methods:** We conducted a systemic search using LitCovid and MEDLINE electronic databases. We screened citations, titles and abstracts, then reviewed potentially relevant articles in full. After data extraction, we reported our final data under subheadings of demographics, diagnosis, clinical presentation, laboratory investigations, imaging studies, treatment, and patient outcomes.

**Results:** Our search strategy yielded 42 original studies reporting 674 pediatric patients fitting the case definition of MIS-C. The studies included 21 case reports, 16 case series and 5 cohort studies. The most common reported symptom of MIS-C was fever (98%). Gastrointestinal symptoms were common (N=557, 83%). Interleukin-6 (IL-6) levels were measured in 125 patients and was elevated in 94 % (N=117). Echocardiography detected coronary artery lesions in 100 patients. Prophylactic and/or therapeutic heparin was required in 34% (N=227) of patients. The most commonly administered treatment modality targeting MIS-C was intravenous immunoglobulin (IVIG) (N=490). Corticosteroids (N=347) and aspirin (N=112) were also integral parts of the treatment regimens. Biologic therapy was integrated into the treatment regimen for 116 patients. Intensive care unit (ICU) admission was alarming (N=478, 71%). 9 fatalities were recorded due to MIS-C

**Conclusions:** We believe MIS-C bears pathophysiological resemblance to the well-known Kawasaki disease but with some key differences highlighted. Understanding those differences will aid our management plan for such patients.

## INTRODUCTION

SARS-CoV-2 is widely considered to be the greatest infectious disease health threat of the 21^st^ century. By the end of June 2020, COVID-19 had already affected more than 10 million people globally. So far, 3 people die every minute because of COVID-19 [1]. The case fatality ratio in adult patients has been reported to be between 1-3% [2]. Unfortunately, data pertaining to pediatric patients is less robust. In addition to lower reported case fatality ratios, pediatric patients appear to have mostly asymptomatic or mild infections [3,4].

Despite having a more favorable clinical course, a global state of vigilance has recently been called for when dealing with pediatric COVID-19 patients. In some of these patients, COVID-19 presented with a severe life-threatening inflammatory syndrome with multiple organ involvement [5]. This newly emerging disease entity was first reported by Jones et al. in May [6]. The yet unnamed syndrome caused confusion due to the overlapping clinical characteristics with Kawasaki disease and toxic shock syndrome. Consequently, the Centers for Disease Control and Prevention (CDC) and the Royal College of Paediatrics and Child Health (RCPCH) published case definitions and recommendations for physicians in managing multisystem inflammatory syndrome in children (MIS-C) [7,8].

These advisory statements have undoubtedly increased case detection. Additionally, as more clinical data is being published about MIS-C, physicians are becoming better at characterizing the syndrome. However, limitations in information still exist as regards the risk factors, pathogenesis, clinical course, and treatment for MIS-C. The primary purpose of this scoping review is to reduce the knowledge gaps and help enable physicians to diagnose and make informed clinical decisions when caring for patients with MIS-C.

## MATERIALS AND METHODS

We included primary studies coinciding with the 2020 COVID-19 pandemic of patients fulfilling the case definition of MIS-C as determined by the CDC [7]: An individual aged %21 years presenting with fever, laboratory evidence of inflammation, and evidence of clinically severe illness requiring hospitalization, with multisystem (>2) organ involvement (cardiac, renal, respiratory, hematologic, gastrointestinal, dermatologic or neurological); AND no alternative plausible diagnoses; AND positive for current or recent SARS-CoV-2 infection by RT-PCR, serology, or antigen test; or exposure to a suspected or confirmed COVID-19 case within the 4 weeks prior to the onset of symptoms.

Our scoping review was designed based on the Arksey and O’Malley framework [9].

### Stage 1: Identifying research questions

Our review of the recently emerging entity of MIS-C in COVID-19 was guided by the following questions; What are the demographics of MIS-C? How is SARS-CoV-2 infection confirmed? What is the clinical presentation of MIS-C? What are the laboratory and imaging characteristics of this disease? What are the possible management strategies? What is its prognosis?

### Stage 2: Identifying relevant studies

We conducted a systematic search on LitCovid and MEDLINE electronic databases; the search was last updated on July 8, 2020. We did not impose any language restriction. Following study selection (stage 3), we hand-searched the reference list of the included studies and additional studies were retrieved. The detailed search strategy could be found in our supplementary material.

### Stage 3: Study selection

Two authors (SH and NF) independently screened citations, titles and abstracts, then reviewed potentially relevant articles in full. This process was repeated independently by two other authors (MK and MS) and results were compared. We considered any article reporting original research on COVID-19 and MIS-C, hyperinflammatory syndrome, or Kawasaki-like illness, whether the diagnosis of COVID-19 was confirmed by RT-PCR, serology or based on clinical, imaging or even through epidemiological contact with confirmed cases. If agreement on abstract or full article inclusion could not be reached between the two reviewers, an opinion was requested from a third reviewer (SE)

A detailed flowchart of the study selection process can be found in figure 1.

### Stage 4: Data charting process

A data-charting electronic form was jointly developed by two reviewers (MK and SE) to extract relevant variables. All authors independently extracted data and continuously updated the data-charting form. We extracted the following data items: general data (title, date of publication, authors’ names, country); methodological data (sample size, participant characteristics - e.g. age, gender and ethnicity); and clinical data (method of COVID-19 confirmation, clinical presentation, laboratory and imaging findings, treatment(s) administered, patient outcomes). As this is a scoping review, we did not perform a formal critical appraisal of primary studies.

### Stage 5: Summarizing results

The results were reported under the following categories: demographics, diagnosis, clinical presentation, laboratory investigations, imaging studies, treatment, and patient outcomes.

We reported the review following the Preferred Reporting Items for Systematic Review and Meta-Analysis (PRISMA) guidelines-extension for scoping review [10].

## RESULTS

Our search strategy yielded 42 original studies reporting 674 pediatric patients fitting the case definition of MIS-C. All studies were published in 2020. Figure 1 shows the study inclusion process in this scoping review. The studies included 21 case reports, 16 case series and 5 cohort studies. The distribution of study designs included in the review are summarized in Table 1.

We included studies from the following countries: 19 from the USA [6,11-28], 7 from the UK [29-35], 6 from France [36-41], 3 from Italy [42-44], 2 from both India [45,46]and Switzerland [47,48], and 1 from each of Luxembourg [49], Israel [50], and Turkey [51]. The records of 8 patients were removed twice because of duplication in 3 studies [31,34,35].

### Demographics

Our study population had a mean age of 8.9 years. There was a slight male predilection (N=381 males vs 292 females). One child did not have their gender specified. The highest reported ethnicity was Afro-Caribbean (25%). Other minor reported ethnicities included ten mixed, three Indian, and two Middle Eastern patients. Out of the total number of patients, 135 were overweight/obese and 95 patients had pre-existing comorbidities: 47 had an unspecified respiratory condition, 23 had bronchial asthma, 11 had autoimmune disease, 7 had cardiac disease, and 7 had other conditions. The demographics are summarized in Table 2.

### Diagnosis

The majority of patients (N=665) underwent reverse transcription-polymerase chain reaction (RT-PCR) testing. RT-PCR had a confirmatory yield of 39% (N=258). Five-hundred and thirty three patients underwent serological testing. It had a confirmatory yield of 86% (N=457). Around 8% of the patient population (N=57) were negative using both testing modalities. Notably, 175 patients had a positive epidemiologic link to a known COVID-19 case. Table 3 shows the results of confirmatory testing.

### Clinical Presentation

The most common reported symptom of MIS-C was fever (98%) lasting between 1 to 19 days. Gastrointestinal symptoms –diarrhea, abdominal pain, and vomiting-were particularly common (N=557, 83%). Furthermore, 98 patients (15%) presented with cervical, mesenteric or mediastinal lymphadenopathy. A few cases reported the following symptoms: edema (N=42), hepatitis (N=26),arthralgia (N=12) and epididymo-orchitis (N=4). Table 4 illustrates the frequency of other clinical features among patients.

### Laboratory Investigations

Patients predominantly demonstrated laboratory findings consistent with an acute inflammatory response including neutrophilia (50%), elevated CRP (78%) and hypoalbuminemia (52%). 125 patients had their interleukin-6 (IL-6) levels measured, it was elevated in 94 % (N=117). Additionally, five patients fulfilled the diagnostic criteria for macrophage activation syndrome.

Myocardial involvement was indicated by elevated troponin levels (55%), and elevated lipase was reported in 11 patients. The laboratory findings are demonstrated in Table 5.

Other hematologic findings reported included lymphocytosis (N=7) and coagulopathy, which was uncommonly reported, with elevated PT (N=27) and elevated aPTT (N=6).

### Imaging Studies

Only 325 patients underwent chest imaging during the course of their illness. Out of these patients, 192 of them demonstrated features of atypical pneumonia in the form of either localized opacities (shadows/nodules) (N=75), ground glass opacities (N=14) or lobar consolidation mainly in the lower and posterior lobes (N=72). Pleural effusion was also noted in 93 patients. Echocardiography detected coronary artery lesions in 100 patients. These lesions were either coronary dilatation (N=40) or coronary aneurysm (N=60). In addition, electrocardiography was performed for 20% of patients. The most common finding was sinus tachycardia. Pelviabdominal imaging (CT and U/S) revealed ascites (N=25) and ileocolitis (N=10) in some patients. Table 6 outlines the imaging modalities and findings reported.

### Treatment

Treatment modalities included supportive measures for the COVID-19 illness in addition to specific measures directed to the MIS-C. The major treatment modalities are represented in figure 2. Prophylactic and/or therapeutic heparin was required in 34 % (N=227) of patients. Supplemental oxygen therapy was required in just 16% of patients. Few patients received medications targeting the causative COVID-19 viral illness. Only 7 patients received hydroxychloroquine and 4 received remdesivir. On the other hand, antibiotics, mainly 3^rd^ generation cephalosporins, were used in 169 patients.

The most commonly administered treatment modality targeting MIS-C was intravenous immunoglobulin (IVIG) (N=490). IVIG resistance was encountered in 11% (N=73) of these patients. Corticosteroids (N=347) and aspirin (N=112) were also integral parts of the treatment regimens. Biologic therapy was integrated into the treatment regimen for 116 patients including anakinra (N=56), infliximab (N=9), and tocilizumab (N=52). Only 2 patients received convalescent plasma.

### Patient Outcomes

Multiorgan system disease was reported in the majority of our included studies. Many patients presenting with MIS-C required intensive care unit (ICU) admission (N=478, 71%). The length of ICU stay ranged from 1 to 46 days with a median of 6 days. The most common reported cause for ICU admission was hemodynamic instability necessitating vasopressor and inotropic support.

Patients required organ support in the form of vasopressors and inotropes (N=377, 56%), invasive mechanical ventilation (N=152, 23%), extracorporeal membrane oxygenation (ECMO) (N=30, 4%), and renal replacement therapy (N=9, 1%). Acalcular cholecystitis was observed in 6 patients. Despite the severity of disease, only 9 fatalities were recorded due to MIS-C; four were due to neurologic events.

## DISCUSSION

A consistent understanding of MIS-C temporally associated with SARS-CoV-2 infection is forming, and the distinction from Kawasaki disease (KD) is emerging. Our scoping review describes the clinical profiles of 674 pediatric patients who met the criteria for this syndrome. Cases have been reported worldwide from over nine countries. Despite underreporting of ethnicities, with around 70% (N=475) of cases having a reported ethnicity, Afro-Carribean and Hispanic ethnicities constituted around half of all cases. This pattern was consistent with findings reported by Feldstein et al. [26]. The mean age was 8.9 years and there was a slight male gender predilection. Conversely, KD tends to affect Asian children younger than 5 years with a similar slight male predominance [52,53]. The presence of comorbidities correlates with severe COVID-19 illness in adults [54]. This correlation may also be consistent with developing MIS-C; 14% of affected children had an underlying illness. However, only 20% had an elevated body mass index which is close to the worldwide prevalence of this condition.

Ecological time-period analyses have verified an association between the sudden rise of Kawasaki-like disease, later termed MIS-C, and the rise of SARS-CoV-2 circulation in a population [25,55]. In fact, Verdoni et al. reported a 30-fold increased incidence of Kawasaki-like disease [43]. Serological testing had a higher confirmatory yield of COVID-19 status (86%) in contrast with RT-PCR (39%). Current understanding of the pathogenesis of this syndrome is that it is a post-infectious sequela triggered by an immune-mediated injury. This may help explain why viral particles are not detected in the majority of patients.

The multiorgan involvement characterizing MIS-C was most frequently manifested by fever (98%), gastrointestinal symptoms (83%) and skin rash (55%). Even though the range of fever spanned 1 to 19 days, we noticed that most patients had a fever duration of 8 days or less. Although clinical signs suggestive of KD -skin rash, mucocutaneous inflammation and cervical lymphadenopathy-were frequent, most patients only fulfilled the criteria for incomplete KD [37,40].Gastrointestinal symptoms were particularly common (83%) in MIS-C. This is in stark contrast with KD which has a lower rate of gastrointestinal involvement [56,57].

Acute kidney injury (AKI) was one of the presenting features in around 16% of patients. Fortunately, only 1% of patients required renal replacement therapy. The incidence of AKI is comparatively low in patients with KD and only a minority require some form of dialysis [58]. Almost one-fifth of patients reported neurological symptoms. Most of these symptoms were minor but Abdel-Mannan et al. suggested that there may be a high incidence of underlying brain MRI signal changes [29].

A hyperinflammatory shock state induced by inflammatory cytokines, which are integral to the pathogenesis of SARS-CoV-2 infection, was typified by 43% and 49% of patients presenting with hypotension and cardiac symptoms, respectively. Most of these patients eventually required vasopressor and/or inotropic support (56%). In contrast, approximately 5% of children with KD in the United States present with cardiovascular collapse and hypotension requiring vasopressor and/or inotropic support [59]. Another interesting finding was that in spite of 49% of patients having initial presentation suggestive of cardiac affection, 55% and 80% eventually developed laboratory and echocardiographic evidence of cardiac pathology during hospitalization, respectively.

Coronary artery abnormalities, an important complication of KD, have been reported in 18% of those who underwent echocardiography. However, only 11% (N=60) of patients developed coronary artery aneurysms (CAA) as opposed to the proposed 25% incidence of CAA in patients with KD [60]. MIS-C patients who developed CAA were generally older and had higher levels of inflammatory cardiac markers when compared to historical KD cohorts [34].

We observed a laboratory pattern highlighting the underlying cytokine storm with elevated CRP, neutrophilia, elevated d-dimer, lymphopenia, and elevated ferritin [61]. When compared to historical cohorts of KD, MIS-C patients were more likely to have neutrophilia, lymphopenia, anemia, elevated CRP, and ferritin [38,34,43]. They also tended to have higher troponin and BNP levels but lower platelet counts. In addition, macrophage activation syndrome, a form of cytokine storm, was reported in 5 patients. Given its role as a marker of severity of COVID-19 illness, hypoalbuminemia was predictably present in 52% of patients [62]. Although data has been published on the use of IL-6 as a prognostic marker in COVID-19, only 19% (N=125) of MIS-C patients underwent interleukin testing [63].

According to the American Heart Association (AHA) guidelines on the treatment of KD, a single infusion of IVIG (2g/kg) should be administered along with aspirin until the patient is afebrile. The addition of glucocorticoids was suggested only for patients at high risk of developing coronary artery aneurysms [64]. However, most centers treating MIS-C cases considered them to be high risk patients, and therefore administered IVIG, aspirin and pulse steroids concurrently. The incidence of IVIG resistance (11%) was similar to the reported rates for KD. Biologic therapy (e.g. anti-IL-6, anti-TNF, etc..) was used independently of IVIG resistance sometimes with the initial regimen on admission. This treatment decision may have been influenced by recent experience with these drugs in adult COVID-19 patients.

Almost a third of MIS-C patients required prophylactic/therapeutic anticoagulation. None of the included studies reported coronary occlusion which is the main indication for anticoagulation in KD [64]. In fact, most studies did not list their indications for anticoagulation. However, it is logical to infer that some of these patients were at particularly high risk of venous thromboembolism given these risk factors: multi-organ system involvement, markedly elevated d-dimers and inflammatory markers, pediatric ICU admission, and the prothrombotic effect of the SARS-CoV-2 infection itself [65].

We found that a significant number of patients required ICU admission (71%). This alarming rate is much higher than rates reported for KD (3%) [66,67]. The benefits of ECMO in adults with COVID-19 are controversial due to the associated risk of thromboembolism especially in those with cytokine storm syndrome [68]. We observed a similar association in our study population; 30 patients ultimately required ECMO, and 7 of them died including 3 by acute neurologic events. However, this association may be confounded by the deteriorating physiological parameters that ultimately led to the need for ECMO.

Our study had several limitations. The first and foremost is that we did not assess the quality of reports. We understand that during a pandemic, the intention to share data rapidly might have an impact on the quality of published primary reports. Some of the primary sources might overlap. We have traced the cases through careful data collection and contacting the authors to minimize the possibility of double counting. Another limitation is that the included studies used variable methods of reporting. We contacted several study authors to obtain primary data and reduce inter-study variability. Several studies had incomplete data for their patients. We understand this may introduce bias in the reporting of certain variables such as mortality. Finally, most studies did not report the exact confirmatory method for diagnosing MIS-C, whether RT-PCR or serology, on an individual basis. We also selected patients according to the CDC case definition only.

In conclusion, we believe MIS-C bears pathophysiological resemblance to the well-known Kawasaki disease. In fact, respiratory viruses, including coronaviridae, have also been implicated in the etio-pathogenesis of KD [69-71]. This review summarizes the major points of difference between both entities. The predominant hyperinflammatory features, the delayed onset after SARS-CoV-2 infection, and the similarity to KD support the hypothesis that MIS-C is a syndrome of immune-mediated injury [72]. This proinflammatory effect of SARS-CoV-2 has also been reported in individuals with severe COVID-19 illness [73,74]. We recommend that future studies are designed to outline prognostic variables and effective treatment strategies. There is still insufficient data to guide cardiac follow-up for MIS-C patients. The use of serological titers for risk stratification is another potential research question [75].

## Data Availability

All data are fully available

## Acknowledgement

We thank Dr. Mostafa El-Hodhod (Prof of Pediatrics. Faculty of Medicine. Ain shams University) for his assistance and his expert opinion on the subject. We thank Mohamed Elshebiny and Farida Elshafeey for their valued contribution to this paper.

